# Development and validation of a dynamic prediction model for unplanned ICU admission and mortality in hospitalized patients

**DOI:** 10.1101/2022.08.30.22279381

**Authors:** Davide Placido, Hans-Christian Thorsen-Meyer, Benjamin Skov Kaas-Hansen, Roc Reguant, Søren Brunak

## Abstract

Frequent assessment of the severity of illness for hospitalized patients is essential in clinical settings to prevent outcomes such as in-hospital mortality and unplanned ICU admission. Classical severity scores have been developed typically using relatively few patient features, especially for intensive care. Recently, deep learning-based models demonstrated better individualized risk assessments compared to classic risk scores such as SOFA and NEWS, thanks to the use of aggregated and more heterogeneous data sources for dynamic risk prediction. We investigated to what extent deep learning methods can capture patterns of longitudinal change in health status using time-stamped data from electronic health records. We used medical history data, biochemical measurements, and the clinical notes from all patients admitted to non-intensive care units in 12 hospitals in Denmark’s Capital Region and Region Zealand during 2011-2016. Data from a total of 852,620 patients and 2,241,849 admissions were used to predict the composite outcome of unplanned ICU transfer and in-hospital death at different time points after admission to general departments. We subsequently examined feature interpretations of the models. The best model used all data modalities with an assessment rate of 6 hours and a prediction window of 14 days, with an AUPRC of 0.287 and AUROC of 0.898. These performances are comparable to the current state of the art and make the model suitable for further prospective validation as a risk assessment tool in a clinical setting.

## Introduction

Early warning scores (EWS) are used in the clinic to assess the health status of hospitalized patients. The first EWS were based solely on five physiological parameters followed by many adaptations and improvements (1), (2). VitalPAC was one such early warning score (ViEWS), which introduced modifications to the parameters contributing to the score (3). These modifications were based on clinicians’ knowledge about the relationship between physiological data and adverse clinical outcomes. Further modifications were implemented in the national early warning score, NEWS (4).

These scores are used extensively because their simplicity makes them applicable and easily comparable across different departments and countries. They specifically provide estimates for the risk of adverse outcomes (5) such as cardiac arrest, ICU transfer, and in-hospital mortality. However, they suffer from some limitations. The relatively few data types used limit their predictive power. Relevant features like biochemical measurements, the order of temporality in the data, or non-linear feature interactions are ignored. These risk calculations are made without exploiting all the information that current EHR systems provide (6). Moreover, the scores mostly rely on data that are manually collected by the staff. This often entails a higher number of missing and incorrect data items, as compared to a fully automated approach. Recently it was shown in population-wide data from inpatients of the Capital Region of Denmark, that around 10% of the NEWS records were incomplete and 0.2% had implausible features (7).

To improve prediction accuracy and reduce alarm fatigue, new applications using more data and more sophisticated methodologies have been developed (8). Given the heterogeneous nature of EHR data and the rarity of events such as clinical deterioration, machine learning methods seem well-suited for addressing this task. Deep learning, in particular, has performed well for many similar clinical tasks. For example, Shamout et al. (9) developed a deep learning model that uses vital signs to predict the composite outcome of ICU transfer, cardiac arrest and mortality. Cho et al. (10) developed a deep learning model to predict the composite outcome of cardiac arrest and ICU transfer, while da Silva et al. (11) developed a long-short term memory neural network (LSTM) to predict the worsening of the vital signs using prognostic indexes. A recurrent deep neural network has also been used by Tóth et al. (12) to predict stability of vital signs to avoid unnecessary measurements at nighttime. Similarly, dynamic deep learning models using multiple data sources have been developed for different tasks; Thorsen-Meyer et al. (13) constructed one to predict 90-days mortality after ICU admission, Rajkomar et al. (14) to predict in-hospital death, 30-day unplanned readmission, prolonged length of stay and patient’s final discharge diagnoses; Tomašev et al. (15) to predict kidney failure; Lauritsen et al. to predict risk of sepsis and acute critical illness (16) (17).

Although deviating vital signs often precipitate clinical deterioration, their recognition in the general ward requires continuous engagement by health care personnel, which can delay or even preclude their recording. Therefore, we investigated using data collected less frequently in early detection of deterioration, building an end-to-end machine learning pipeline that integrates heterogeneous clinical data to predict the risk of imminent serious clinical deterioration at regular intervals.

To this end, we combined natural language processing algorithms and recurrent neural networks to leverage latent patterns in the data. We subsequently assessed the impact of the single data sources and the so-called tokens extracted from each of them to improve the understanding of the model.

## Methods

This paper adheres to relevant items in the Transparent Reporting of a multivariable prediction model for Individual Prognosis or Diagnosis statement (TRIPOD) (18).

### Patients and outcome

The data comprise all inpatient admissions, from 2011 to 2016, to 12 public hospitals in the Capital Region of Denmark and Region Zealand. The admissions were pieced together by concatenating consecutive inpatient visits 24 hours apart so that the department transfers were not considered two separate events.

Direct ICU admissions, outpatients and emergency admissions were excluded, as were individuals with disconnected medical record history (either because these patients moved to another country or lacked a stable residence) and minors (age <16 years).

The outcome *clinical deterioration* was defined as unplanned ICU transfer or in-hospital mortality within the so-called prediction window. Unplanned ICU transfer was defined as any acute admission to an ICU (filtering by the codes NABE and NABB from the Sundhedsvæsenets Klassifikations System (SKS) - the Danish Health Service Classification System) within 24 hours after discharge from a non-ICU ward.

### Model

The model architecture (Figure2) was designed as a scalable network that can be adapted to different data modalities by using one sub-model per data domain; each sub-model consists of an embedding layer (which transforms the categorical variables into vectors), a recurrent neural network (which learns from the sequence of embedding vectors) and a pooling layer (which reduces the vectors’ dimensionality).

**Figure1:**
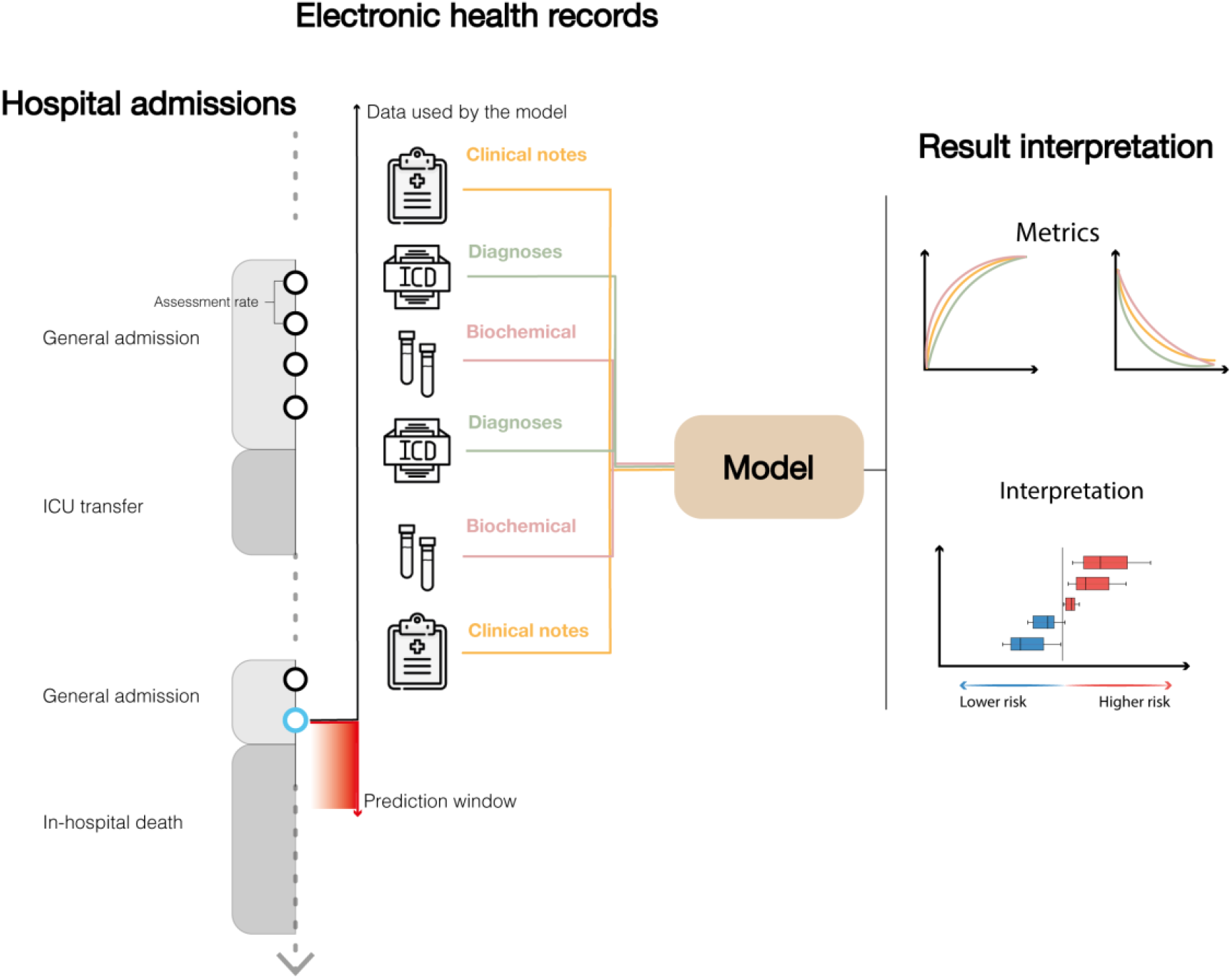
General structure of the prediction framework. Given a specific assessment rate (time between two consecutive risk assessments) and prediction window (time window within which the outcome is observed), the risk of clinical deterioration was assessed continuously during each general admission. All the data up to the time of assessment was used to train the model, which was then evaluated and interpreted on the holdout test data.

**Figure2:**
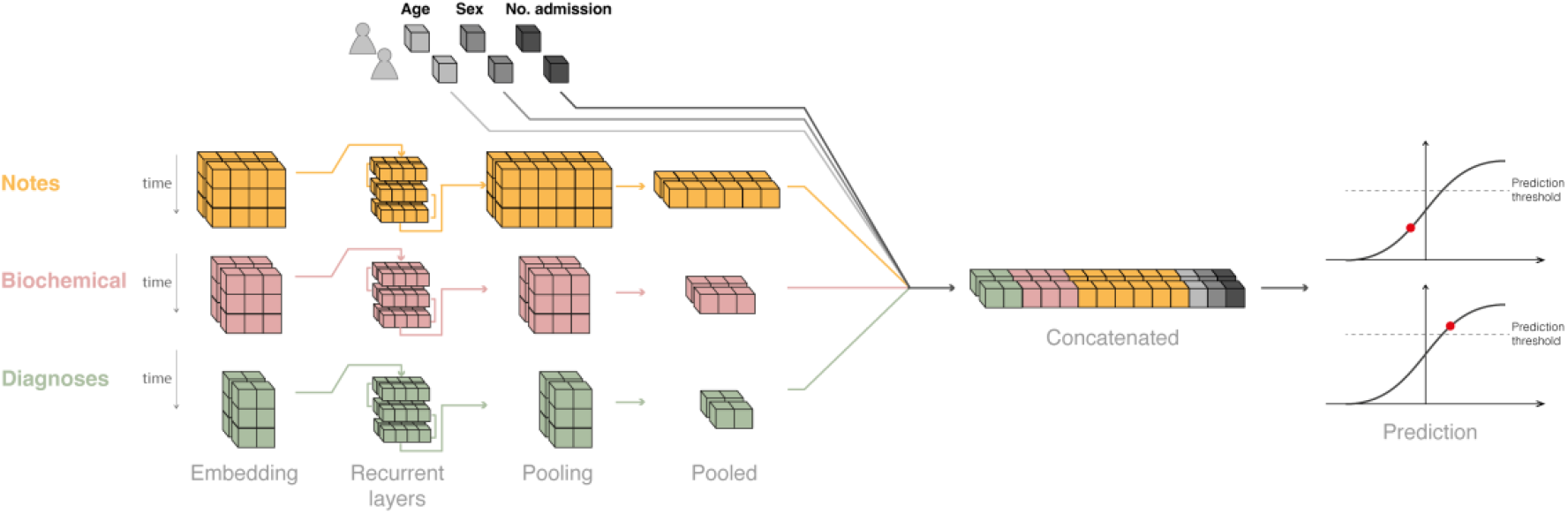
Structure of the deep learning model, exemplified by two admissions. For each sample (i.e. admission at a specific time point) a 2D tensor (matrix) comprises the sequence of embedded tokens until the time of assessment. Each tensor was given as input to a recurrent block (GRU or LSTM, part of the hyperparameters search) and the time dimension was pooled through an attention pooling layer. The flattened tensors were then concatenated and fed to a linear layer with a standard logistic activation function.

The network uses tokens, i.e. sequences of characters grouped together based on their semantics, as input. We chose the entity embedding approach to exploit the heterogeneity and sparseness of the input data, thus mapping the tokens constructed from the categorical features to embeddings in a Euclidean space (i.e. the embedding space) (19). For each sub-model, the vocabulary size V (i.e. the number of unique tokens for the specific data source) was directly used to calculate the size of the embedding representation according to 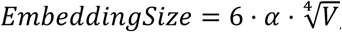, where the embedding coefficient *α* was estimated by hyperparameter search (14).

We explored two types of recurrent neural networks, Gated Recurrent Unit (GRU) (20) and Long Short-Term Memory (LSTM) networks (21). Rather than max or average pooling – which retains neither positional nor intensity information – we used an attention-based pooling (22) which employs a weighted mechanism to retain the most relevant parts of a sequence. The attention-based pooling layer was used to aggregate the temporal dimension of the recurrent layer output, which was then concatenated across the different data sources. The final layer was a linear (= dense) layer with a standard logistic activation function hence mapping the output into the interval [0, 1] to yield valid predicted probabilities. The features without temporal components (age, number of previous hospital admissions and sex) were fed into the model by concatenating them to the output of the pooling layer.

### Data sources and processing

The input data comprised medical disease history, biochemical measurements, clinical notes and demographics (age, sex and number of previous admissions). The medical disease history prior to the start of the EHR data was also used (as well as after to handle false negative outcomes not covered after the EHR data period). A schematic of the time spans for the different data modalities is depicted in FigureS1. The disease history was extracted from the Danish National Patient Registry (DNPR) for the period between 1977 and 2018 (23). DNPR is nation-wide registry which covers essentially all the hospital encounters in Denmark. The disease codes use Danish adaptations from SKS of the International Classification of Diseases (ICD) version 8 up to 1993, and ICD-10 from 1994 and onwards. Only diagnosis codes recorded prior to the admission date were included in the predictive schemes.

Biochemical values were extracted from the Clinical Laboratory Information System (LABKA) and the Clinical Chemistry Laboratory System (BCC) databases (24) for the period overlapping the hospital admissions (2011-2016). These databases collect all the biochemical tests performed by the Danish hospital laboratories in the Capital Region and Region Zealand, respectively. The biochemistry tokens were constructed using the name of the biochemical component, the specimen (blood, plasma etc.), the unit and the quantile of the value of the measurement (to yield a vocabulary with a reasonable size); for example, *HEMOGLOBIN_B_mmol/L@8-8.5*. The quantile binning was included as a hyperparameter.

The clinical notes were extracted from the EHR data for the admissions from 2011 to 2016. The free text required some preprocessing before tokenization, such as removing punctuation, names, stop words (25), negations and signatures of the clinicians. Due to the large number of terms coming from the medical notes, the embedding of this specific data source was trained separately on the full corpus using fastText (26), to reduce the number of parameters to update during training.

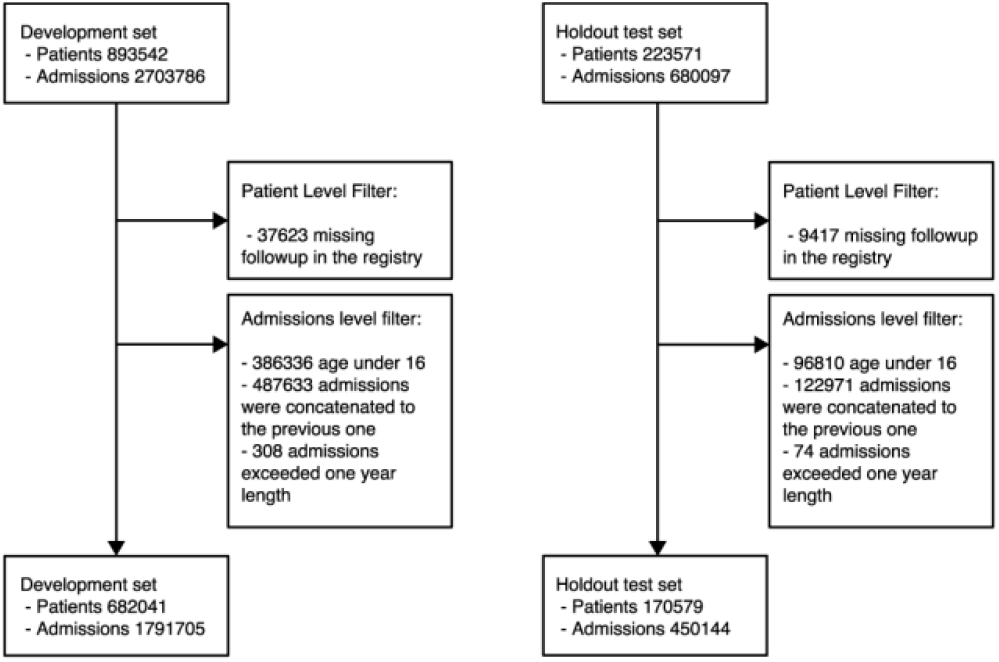
Attrition diagram: Study profile.

### Training and Evaluation

We randomly split the dataset into a development set (80%) for model creation and an independent holdout test set (20%) for model evaluation. The split of the dataset was done at the patient level and assigned admissions of the same patient to the same set to avoid leaking information between the sets (27). The development set was further split into a training set (80%, 64% of total) and validation set (20%, 16% of total) to counter overfitting and to calibrate the model before testing it on the holdout set.

Three submodels were trained separately to find the best submodel-specific hyperparameters; we used Optuna’s multivariate TPEsampler, based on the Three-structured Parzen Estimator (TPE) algorithm to search the hyperparameter spaces (28,29). While searching the hyperparameter space, we fixed the prediction window at 24 hours and the assessment rate at 12 hours, to facilitate the comparison across the different data types. TableS1 shows the hyperparameters explored for each submodel; FiguresS2-S4 illustrate the hyperparameter searches. For each experiment, the loss on the validation set was used as training metric to select the model at the best epoch. The performance metric used to select the best model for each search was the area under the precision-recall curve (AUPRC). Although the area under the receiver operating curve (AUROC) score is a more common metric for classification tasks, in this case it would not be sufficient to appreciate the real ability of the model to discriminate between the two classes due to their considerable imbalance (30). AUPRC on the other hand is much more robust to class imbalances. We used 200 bootstraps samples (31) to construct the 95% confidence interval for the metrics shown in figures and tables.

Using the best submodel architectures, we trained and evaluated an ensemble model for different prediction windows (1, 2, 7, 14 days) and assessment rates (every 6, 12 and 24 hours). Finally, we applied post-hoc isotonic calibration to align the final predicted risk with the actual outcome incidence, using the data in the validation set (32). The model re-calibration was achieved by fitting an isotonic regression using the output of the model prior to calibration as regressor and the actual label of the samples as response variable. We used the validation set to fit the isotonic regression and kept the test set untouched. The fitted isotonic model was used to adjust the output of the uncalibrated model to get calibrated predictions.

All the results reported are generated using the re-calibrated model on the test set. To control for biases driven by age or sex, performances were also evaluated at each time of assessment for the different subgroups.

### Interpretation

The impact of the different tokens on the model outcome was calculated using the GradientShap algorithm (33) from the *Captum* library (34). Given an input feature of a single risk assessment, its Shap value is correlated to how much (and in what direction) that feature pulls the individual-level prediction away from the population-level mean risk. Importantly, Shap values do not represent the effect of a single feature on the model outcome but rather the effect of that feature in the context of a coalition of features. Shap values were calculated for the best model after isotonic re-calibration.

## Results

The model was trained on 682,041 unique patients and 1,791,705 admissions and evaluated on 170,579 patients and 450,144 admissions as described in Methods. In both parts, 1.5% of the admissions resulted in in-hospital mortality and 0.7% in ICU transfer. 2,583 tokens from the ICD code data type (medical history), 2,421 tokens from the biochemical measurements and 403,869 tokens from the medical notes were used as input. We investigated the inclusion of each data type separately as well as jointly in the same model.

When we explored how the prediction window and assessment rate affect the performances, the most performant model was the one trained on all the data sources using a prediction window of 14 days and an assessment rate of 6 hours with an AUROC of 0.904 [0.903-0.905] and AUPRC 0.285 [0.283-0.287] over all the predictions (Table2). This model was after isotonic recalibration well-calibrated (FigureS5) with a calibration slope of 0.964 [0.951, 0.98], an intercept of 0.002 [0.001-0.003] and an upper bound risk of 79%.

**Table1.** Summary statistics for the Development and Test sets. Statistics are calculated at the admission level.

|  | Development Set | Test Set |
| --- | --- | --- |
| Number of Patients | 682041 | 170579 |
| Number of Admissions | 1791705 | 450144 |
| Age at prediction<br>- Discharge<br>- ICU Transfer<br>- In-hospital Death | 60 [40-73]<br>67 [56-75]<br>78 [68-86] | 60 [40-73]<br>67 [56-75]<br>78 [69-86] |
| Sex:<br>- Male<br>- Female | 798185 (45%)<br>993520 (55%) | 199732 (44%)<br>250412 (56%) |
| Outcome<br>- Discharge<br>- ICU Transfer<br>- In-hospital Death | 1752329 (97.8%)<br>13129 (0.7%)<br>26247 (1.5%) | 440221 (97.8%)<br>3305 (0.7%)<br>6618 (1.5%) |
| Number of previous Admissions<br>- Discharge<br>- ICU Transfer<br>- In-hospital Death | 1 [0-4]<br>2 [0-6]<br>3 [1-7] | 1 [0-4]<br>2 [0-6]<br>3 [1-6] |
| Length of stay before outcome (Hours):<br>- Discharge<br>- ICU Transfer<br>- In-hospital Death | 31 [9-98]<br>35 [8-121]<br>126 [43-285] | 31 [9-97]<br>35 [8-132]<br>122 [43-279] |
| Type ICU Transfer<br>- Surgical<br>- Medical | 2358 (0.18%)<br>10771 (0.82%) | 639 (0.20%)<br>2666 (0.80%) |

**Table2.** Performance on the test set for models using all data types.

| Frequency of assessment | Prediction window | AUROC | AUPRC | Calibration |  |
| --- | --- | --- | --- | --- | --- |
|  |  |  |  | Slope | Intercept |
| 6h | 1d | 0.925 [0.923-0.926] | 0.152 [0.149-0.155] | 1.042 [1.02-1.064] | -0.001 [-0.001-0.0] |
|  | 2d | 0.922 [0.921-0.923] | 0.180 [0.177-0.184] | 0.988 [0.966-1.007] | 0.002 [0.001-0.003] |
|  | 7d | 0.913 [0.912-0.914] | 0.269 [0.267-0.271] | 0.996 [0.985-1.008] | 0.002 [0.001-0.003] |
|  | 14d | 0.904 [0.903-0.904] | 0.285 [0.283-0.286] | 0.964 [0.951-0.98] | 0.004 [0.003-0.005] |
| 12h | 1d | 0.918 [0.916-0.920] | 0.129 [0.125-0.134] | 0.998 [0.96-1.045] | 0.001 [0.0-0.002] |
|  | 2d | 0.916 [0.914-0.917] | 0.187 [0.183-0.191] | 0.996 [0.977-1.021] | 0.002 [0.0-0.003] |
|  | 7d | 0.898 [0.897-0.899] | 0.233 [0.231-0.236] | 1.045 [1.026-1.067] | -0.001 [-0.002-0.0] |
|  | 14d | 0.901 [0.900-0.902] | 0.278 [0.275-0.282] | 0.952 [0.912-0.997] | 0.005 [0.001-0.007] |
| 24h | 1d | 0.905 [0.901-0.908] | 0.135 [0.130-0.140] | 1.038 [0.992-1.078] | 0.0 [-0.001-0.001] |
|  | 2d | 0.901 [0.899-0.903] | 0.163 [0.157-0.169] | 1.017 [0.969-1.059] | 0.0 [-0.001-0.002] |
|  | 7d | 0.896 [0.894-0.897] | 0.232 [0.227-0.236] | 1.014 [0.969-1.05] | 0.001 [-0.001-0.003] |
|  | 14d | 0.893 [0.892-0.894] | 0.250 [0.246-0.254] | 0.974 [0.951-1.006] | 0.003 [0.001-0.005] |

The performances were similar across sexes, with AUPRCs of 0.288 [0.285-0.290] and 0.280 [0.277-0.283] for males and females, respectively. The performances for the different age groups varied more, with a trend of AUPRC increasing with age (age 16-37: 0.168 [0.157-0.180], age 37-58: 0.252 [0.246-0.257], age 58-79: 0.277 [0.274-0.280], age 79-100: 0.310 [0.307-0.313]).

We observed an increased AUPRC for risk estimates further into the hospital stay with a peak at 7 days into the admission FigureS6-S7.

Among the models trained separately using a 24-hour prediction window and 12 hours assessment rate, the most performant was the one trained on the clinical notes with an AUPRC of 0.125 [0.120-0.130]; this value improved to AUPRC of 0.131 [0.127-0.135] when the model was trained on all the data using the same prediction window and assessment rate.

The best encoding of disease diagnoses was rolling diagnoses up to the third ICD level (e.g. C341M to C34) and a padding size of 9 (FigureS2). The optimal quantile resolution for the biochemical data was deciles (i.e. 10 bins) and a padding size of 28 was the optimal number of lab values to include before time of prediction (FigureS3). The optimal padding size for medical notes was 299, reflecting the larger amount of information usually held by the clinical notes (FigureS4). The complete list of the optimal parameters for each search can be found in the supplementary TableS1.

### Feature importance

The importance of the tokens from the different vocabularies can be sorted according to their attributed Shap values (Figure4). Higher age and male sex were associated with elevated risk of deterioration. The opposite was the case for the number of previous admissions: a higher number of previous admissions was associated with lower risk of deterioration (Figure4.A-C).

**Figure3.**
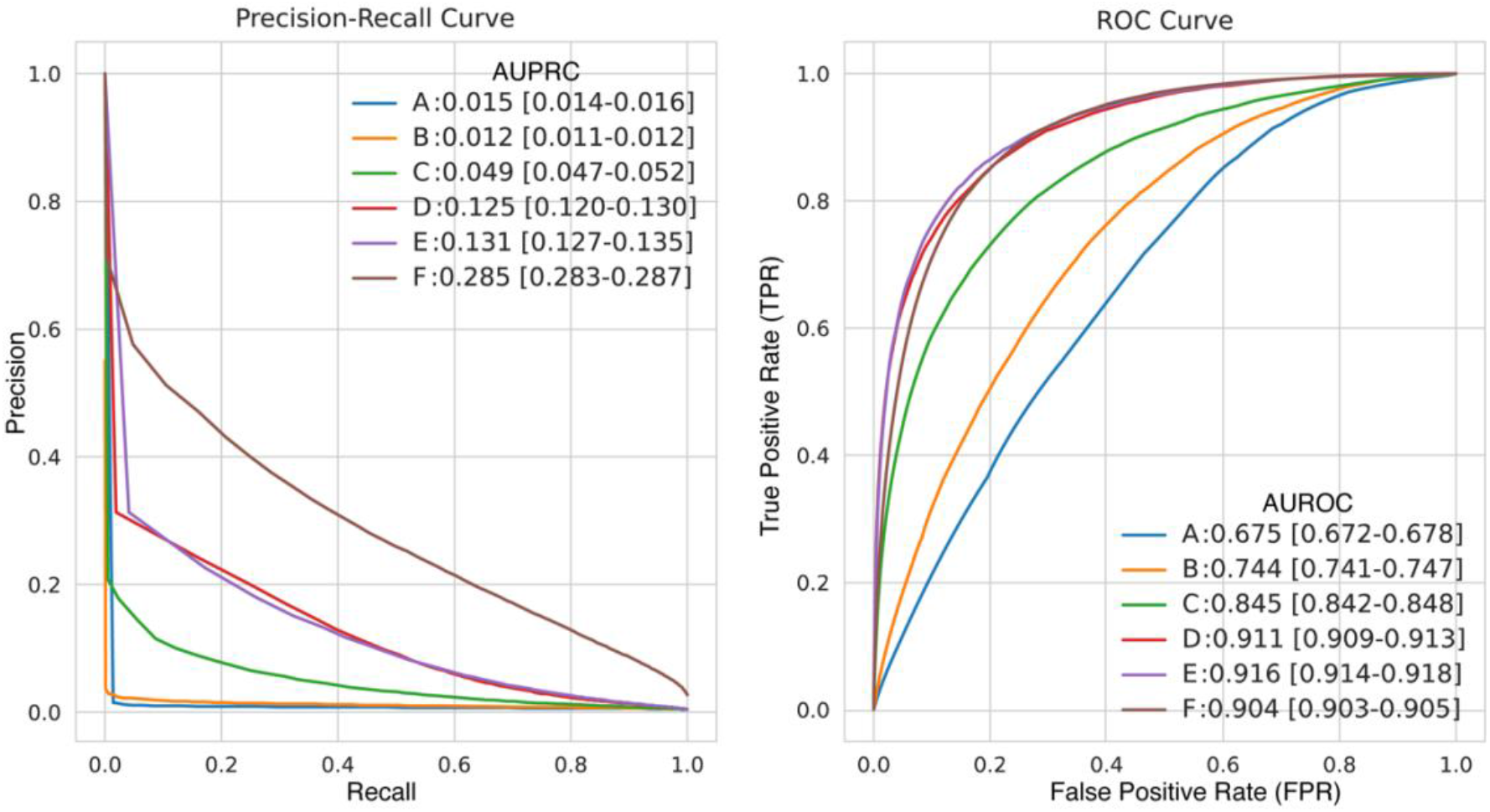
Performance of the model for prediction of unplanned ICU transfer or in-hospital death. Panel A: precision recall curves for the six models. Panel B: receiver operating characteristic (ROC) curves show the values of true positive rate (= recall = sensitivity) and false positive rate (= 1-specificity) at different thresholds for the six models. Model A: age, sex, number of admissions. Model B: model A + medical history data. Model C: model A + biochemical data. Model D: model A + clinical notes. Models E and F: model A + medical history data, biochemical data and clinical notes. Models A–E use a 24-hour prediction window and a 12-hour assessment rate. Model F uses a 14-day prediction window and a 6-hour assessment rate.

**Figure4:**
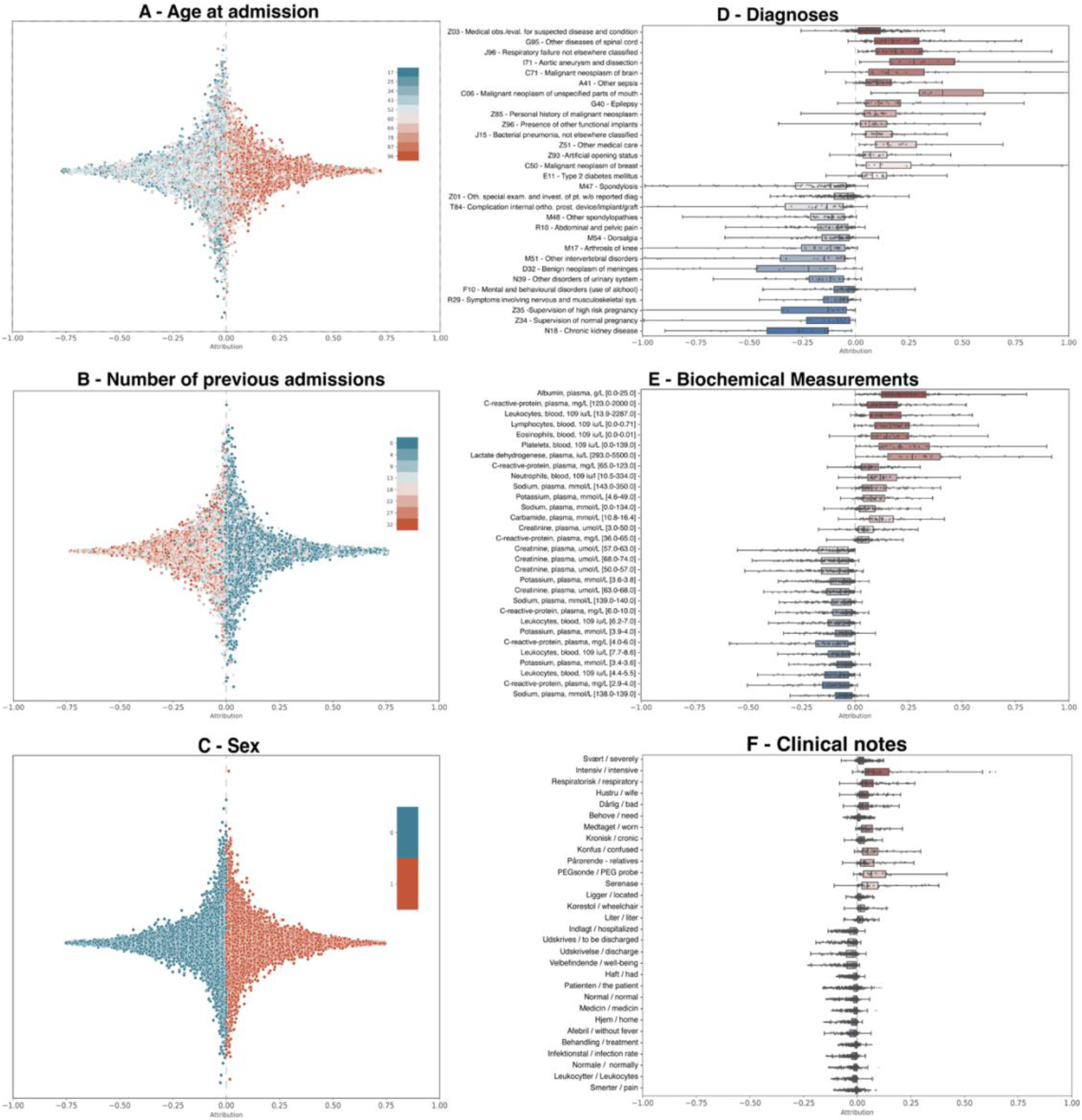
Contribution of the different tokens to the outcome of clinical deterioration. Shap values were estimated using the best model (model using age, sex, number of admissions, medical history data, biochemical data and clinical notes with a 14-day prediction window and 6-hours assessment rate). Panels A-C: the distribution of Shap values for each feature without temporal components, with the color scale representing the feature value. Panels D-F: the distribution of Shap values for top-15 tokens (in up- and downward directions, respectively). Overlain boxes-and-whiskers show medians, quartiles and 1.5 x quartiles.

The diagnosis tokens associated with clinical deterioration were often those of acute illnesses such as respiratory failure, neoplasm and pneumonia (Figure4.D). In contrast, diagnosis codes related to pregnancy, chronic conditions and orthopedics were associated with lower risk.

Low levels of albumin, lymphocytes and sodium were associated with elevated risk of clinical deterioration; the same were high levels of C-reactive protein, leukocytes, sodium, lactate dehydrogenase, potassium and carbamide (Figure4.E). In contrast, normal levels of leucocytes, hemoglobin, C-reactive protein, sodium and potassium were all associated with low risk of deterioration.

The tokens from clinical notes most strongly associated with clinical deterioration were *severely, intensive, respiratory and chronic. Pain, leukocytes, control, normal, home, discharge(d)* and on the other hand, were all associated with low risk (Figure4.F).

## Discussion

The main aim of this study was to explore whether data types registered routinely in general departments are predictive of clinical deterioration, ultimately to assess if these suffice for this task. A solution based on data collected routinely might circumvent a common weakness of EWS, i.e. they depend on data that require clinical engagement (e.g., vital signs). Consequently, offering a viable alternative for risk stratification with minimum additional manual data collection effort is preferable.

Exploiting the combined power of entity embedding of tokens from electronic health records and the ability of recurrent neural networks to learn temporal patterns from such data, we built a performant (AUROC and AUPRC up to 0.90 and 0.28, respectively) and well-calibrated deep learning model for predicting the risk of clinical deterioration. Specifically, leveraging medical history data (up to 40 years) from a national register along with in-hospital biochemical data and clinical notes, we trained the model dynamically, meaning that the same model can handle different time points for the same admission.

Direct comparison of our model to the performances of h is not possible, since the vital signs used for the calculation of such scores are not collected in our EHR dataset. Comparison of the performance of our model to other work is also non-trivial: AUROC is the most common metric for risk stratification and usually the one used to compare performance across studies. Nevertheless, it is unsuited for imbalanced prediction problems (such as the one defined here) because it disregards the prevalence of the outcome of interest (30). Although AUPRC does account for prevalence, it is not always reported in the classic studies on EWS scores. Moreover, direct comparison of the AUPRCs requires equal (or at least similar) prevalence of the outcome across studies; this is problematic because the prevalence tends to vary between cohorts, and, more importantly, so do the criteria used to define it. For example, Watkinson et al. (35) defined the outcome as the composite of ICU transfer, in-hospital death and cardiac arrest with a prediction window of 24 hours (AUROC of 0.868 [0.864-0.872]). Dziadzko et al. (36) defined the outcome as in-hospital death or respiratory failure, with a prediction window of 48 hours (AUROC 0.87 [0.85–0.88] and 0.90 [0.84–0.95] in 2013 and 2017, respectively). Malycha et al. (37) defined the outcome as in-hospital death and ICU transfer within 24 h following assessment from patients admitted longer than 24 hours (AUROC 0.823 [0.819–0.824]). Cho et al. (10) instead defined the outcome as cardiac arrest and unexpected ICU admission, occurring within 0.5-25 hours from the assessment (AUROC 0.865 [CI not reported]). The performance difference of models A-E (Figure3) is driven by information learnt from different data types. The performance does not increase linearly with the addition of new data types, suggesting substantial overlap in the latent information of diagnosis codes, biochemical data and clinical notes. This corresponds well to what one would expect, e.g. a clinical note may very well cite lab values (and likely those of greatest clinical interest) and summarize anamnestic information such as comorbidities also registered in the diagnosis code data.

On the other hand, the change in performance from model E to model F (AUPRC from 0.131 [0.127-0.137] to 0.287 [0.284-0.289]) is likely driven by the different incidence of the outcome when using longer prediction windows, essentially making the prediction task easier because there are more examples to learn from. Also, it is much more difficult to discriminate between patients having a severe outcome in the subsequent 24 or 48 hours, because drivers of short-term mortality/ICU transfer depend not only on the physiological status of the patient but also on many factors beyond what is captured in clinical data, e.g. the coordination of the resources within and between hospital departments. The assessment rates employed are consistent with how often new observations and information are recorded in patient files: new clinical notes and biochemical measurements are usually recorded a couple of times every day and normally at least once. The change in AUPRC and AUROC during the admissions reflects the increasing availability of new records when the patients proceed further into the hospitalization (FigureS6-S7), and that patients tend to deteriorate either early or late in the admission and less in between.

We decided to keep the outcome definition broad rather than predicting specific severe conditions such as sepsis or organ failure, notoriously difficult to operationalize for prediction tasks (38). In contrast, using a general definition of clinical deterioration allowed us to keep the feature space more generic and less dependent on specific illnesses. Indeed, our model seeks not to advice on interventions but to flag patients at risk of (more of less imminent) clinical deterioration so health care staff can intervene in a manner appropriate for the patient in question, hopefully translating into improved prognosis for that patient.

An ensemble structure of the network was preferred over a structure in which the different data types contributed to the creation of the same embedding space. Separate submodels (embedding + recurrent linear layer + pooling) allowed us to tune their architectures; optimal padding sizes and embedding coefficients, for example, differ for diagnosis codes and biochemical data. Its ensemble nature also renders the model scalable for incorporating new data types/domains.

To obtain a reasonably well-defined training cohort, we only included in-patient admissions to general departments, excluding out-patients and acute admissions to the emergency department. The former groups were excluded because they are more unlikely to experience the outcome, the latter because the rapid course of events and acute physiology recorded in the emergency department would necessitate a different experimental setup.

### Interpretation

Although ranking the features that drive the predicted risks up or down is useful as a sanity check of the signals picked up by the model, these estimates are not causal. Indeed, the attribution of each token is determined by its context therefore the same token could be associated with both elevated and diminished risk depending on the other tokens with which it co-occurs. For example, a lab value outside the physiological range (e.g., low creatinine), which would normally drive the risk up, may not affect the predicted risk when co-occurring diagnosis codes counter its contribution (e.g., pregnancy codes).

This is more evident for the free text, where the semantics of a word is always dependent on its context. Overall, the feature attribution does not contain any counterintuitive explanation and the few seemingly questionable interpretations have plausible explanations. For example, we find low eosinophil counts among the lab values associated with elevated risk of clinical deterioration. Clinically, this seems counterintuitive since low values of eosinophils represent the standard and high eosinophil counts are indicative of infections (especially parasitic) and allergic disorders. Eosinophils, however, are part of standard panels for blood differential counts (a count for the different types of white blood cells) and, as such, the very presence of (any) eosinophil count probably reflects a clinical suspicion of infection which caused additional analysis to investigate on the infectious agent. Interpretation of tokens from the clinical notes also provides some examples that at first seem odd but probably do have some contextual bearing. For example, the tokens *wife* (hustru) and *relatives* (pårørende) are strongly associated with clinical deterioration. This is likely because doctors document in the patient file when relatives have been informed or consulted, and this may well be on poor prognoses or even no-resuscitation orders in which case clinical deterioration is almost certain to ensue.

Numerical features like the number of previous admissions should also be contextualized. While higher age is indeed associated with clinical deterioration, having a lot of admissions prior to the one of the assessments does not. This seeming paradoxical result can be probably explained by the ability of the model to integrate data from different EHR domains to recognize patients with chronic conditions who will have more frequent hospital visits but perhaps be less likely to suddenly fall critically ill.

### Strengths

This study has some important strengths. First, it is one of the largest of its kind, with a total of 852,620 patients and 2,241,849 admissions taking place over 6 years. Second, we provided a dynamic risk assessment, showing how prediction performance changes over the course of an admission for different prediction windows. This supersedes early warning scores, based on static metrics that do not take sequential information into account but use just a snapshot of the patient’s current status. Third, thanks to the model’s architecture, adding new data sources is relatively easy. New features can be added and removed from the model, adapting the tool to the available resources. However, as expected, the model performs best with as many data as possible included. Finally, while classic EWS depend on complete data to be calculated, missing data in this setup is not a problem since the model has been designed to handle it thanks to the entity embeddings.

## Limitations

Like any study this has limitations. First, although we tried to define the outcome robustly, there are some pitfalls to consider for ICU transfer and in-hospital mortality. Patients in very severe conditions may still be discharged by the hospital if the latter is not able to provide any support to the patient. These patients will probably experience clinical deterioration within the prediction window but not within their hospital stay, hence they are labelled as negative but potentially still flagged by the model as high-risk patients; this could inflate the number of false positive patients detected by the model. Second, unplanned ICU transfer was captured only for the patients who were admitted in one of the ICUs in our catchment area. It cannot be excluded that some patients admitted to a general department of one of the hospitals in our catchment area are transferred to ICUs of other Danish regions than included here, even if it is uncommon. Third, model performance may improve with additional data such as genetic data, vital signs and other biomarkers. Thanks to ensemble nature, adding such data in setting where they are available is simple. Finally, the model was trained on data from the Danish healthcare system and the model (due to the entity embeddings) would need to be trained anew if deployed in other geographical or healthcare systems.

## Conclusion

Combining entity embeddings and recurrent neural networks we built a highly performant model for flagging patients at risk of clinical deterioration. The model was developed and evaluated in a controlled in-silicon setting. Although the data used were collected prospectively, a proper prospective evaluation would be needed to establish whether its deployment can produce real-world benefits to patients on hard endpoints.

## Data Availability

The authors do not have permission to share data access to the original data can be obtained from the Danish health authorities.

## Data and software statement

The software used in the study is based on python v3.8 and pytorch. The software is available online at https://gitfront.io/r/daplaci/uA6jdq4FtTJQ/ClinicalDeterioration/. The authors do not have permission to share data; access to the original data can be obtained from the Danish health authorities.

## Funding and permission statement

This study was approved by the Danish Patient Safety Authority (3-3013-1731 and 3– 3013–1723), the Danish Data Protection Agency (DT SUND 2016–48, 2016–50, 2017–57 and UCPH 514-0255/18-3000:) and the Danish Health Data Authority (FSEID 00003092, FSEID 00003724, FSEID 00004758 and FSEID 00005191). The Novo Nordisk Foundation (grants NNF17OC0027594 and NNF14CC0001) and the Danish Innovation Found (5184-00102B) supported this study.

## References

1. Smith MEB, Chiovaro JC, O’Neil M, Kansagara D, Quiñones AR, Freeman M, et al. Early warning system scores for clinical deterioration in hospitalized patients: A systematic review. Vol. 11, Annals of the American Thoracic Society. American Thoracic Society; 2014. p. 1454–65.

2. Subbe CP, Kruger M, Rutherford P, Gemmel L. Validation of a modified Early Warning Score in medical admissions. QJM: An International Journal of Medicine. 2001 Oct 1;94(10):521–6.

3. Prytherch DR, Smith GB, Schmidt PE, Featherstone PI. ViEWS—Towards a national early warning score for detecting adult inpatient deterioration. Resuscitation. 2010 Aug 1;81(8):932–7.

4. Jones M. NEWSDIG: The National Early Warning Score Development and Implementation Group. Clinical Medicine. 2012 Dec 1;12(6):501.

5. Churpek MM, Yuen TC, Edelson DP. Predicting clinical deterioration in the hospital: The impact of outcome selection. Resuscitation. 2013 May 1;84(5):564–8.

6. Downey CL, Tahir W, Randell R, Brown JM, Jayne DG. Strengths and limitations of early warning scores: A systematic review and narrative synthesis. Vol. 76, International Journal of Nursing Studies. Elsevier Ltd; 2017. p. 106–19.

7. Pedersen NE, Lars ·, Rasmussen S, John ·, Petersen A, Thomas ·, et al. A critical assessment of early warning score records in 168,000 patients. J Clin Monit Comput. 2018;32:109–16.

8. Al-Shwaheen TI, Moghbel M, Hau YW, Ooi CY. Use of learning approaches to predict clinical deterioration in patients based on various variables: a review of the literature. Artificial Intelligence Review 2021. 2021 Mar 13;1–30.

9. Shamout FE, Zhu T, Sharma P, Watkinson PJ, Clifton DA. Deep Interpretable Early Warning System for the Detection of Clinical Deterioration. IEEE J Biomed Health Inform. 2020 Feb 1;24(2):437–46.

10. Cho KJ, Kwon O, Kwon JM, Lee Y, Park H, Jeon KH, et al. Detecting patient deterioration using artificial intelligence in a rapid response system. Crit Care Med. 2020;E285–9.

11. da Silva DB, Schmidt D, da Costa CA, da Rosa Righi R, Eskofier B. DeepSigns: A predictive model based on Deep Learning for the early detection of patient health deterioration. Expert Syst Appl. 2021 Mar 1;165:113905with.

12. Tóth V, Meytlis M, Barnaby DP, Bock KR, Oppenheim MI, Al-Abed Y, et al. Let Sleeping Patients Lie, avoiding unnecessary overnight vitals monitoring using a clinically based deep-learning model. npj Digital Medicine 2020 3:1. 2020 Nov 13;3(1):1–9.

13. Thorsen-Meyer HC, Nielsen AB, Nielsen AP, Kaas-Hansen BS, Toft P, Schierbeck J, et al. Dynamic and explainable machine learning prediction of mortality in patients in the intensive care unit: a retrospective study of high-frequency data in electronic patient records. Lancet Digit Health. 2020 Apr 1;2(4):e179–91.

14. Rajkomar A, Oren E, Chen K, Dai AM, Hajaj N, Hardt M, et al. Scalable and accurate deep learning with electronic health records. npj Digital Medicine 2018 1:1 [Internet]. 2018 May 8 [cited 2021 Sep 20];1(1):1–10. Available from: https://www.nature.com/articles/s41746-018-0029-1

15. Tomašev N, Glorot X, Rae JW, Zielinski M, Askham H, Saraiva A, et al. A clinically applicable approach to continuous prediction of future acute kidney injury. Nature 2019 572:7767. 2019 Jul 31;572(7767):116–9.

16. Lauritsen SM, Kalør ME, Kongsgaard EL, Lauritsen KM, Jørgensen MJ, Lange J, et al. Early detection of sepsis utilizing deep learning on electronic health record event sequences. Artif Intell Med. 2020 Apr 1;104:101820.

17. Lauritsen SM, Kristensen M, Olsen MV, Larsen MS, Lauritsen KM, Jørgensen MJ, et al. Explainable artificial intelligence model to predict acute critical illness from electronic health records. Nature Communications 2020 11:1. 2020 Jul 31;11(1):1–11.

18. Collins GS, Reitsma JB, Altman DG, Moons KGM. Transparent reporting of a multivariable prediction model for individual prognosis or diagnosis (TRIPOD): The TRIPOD Statement. BMC Med. 2015 Jan 6;13(1):1–10.

19. Guo C, Berkhahn F. Entity Embeddings of Categorical Variables. 2016 Apr 22; Available from: https://arxiv.org/abs/1604.06737v1

20. Cho K, Van Merriënboer B, Bahdanau D, Bengio Y. On the Properties of Neural Machine Translation: Encoder-Decoder Approaches.

21. Gers FA, Schmidhuber J, Cummins F. Learning to forget: Continual prediction with LSTM. IEE Conference Publication. 1999;2(470):850–5.

22. Lin Z, Feng M, dos Santos CN, Yu M, Xiang B, Zhou B, et al. A Structured Self-attentive Sentence Embedding. 5th International Conference on Learning Representations, ICLR 2017 - Conference Track Proceedings [Internet]. 2017 Mar 9 [cited 2022 Aug 25]; Available from: https://arxiv.org/abs/1703.03130v1

23. Schmidt M, Schmidt SAJ, Sandegaard JL, Ehrenstein V, Pedersen L, Sørensen HT. The Danish National Patient Registry: a review of content, data quality, and research potential. Clin Epidemiol. 2015 Nov 17;7:449.

24. Grann A, Erichsen R, Nielsen A, Frøslev T, Thomsen R. Existing data sources for clinical epidemiology: The clinical laboratory information system (LABKA) research database at Aarhus University, Denmark. Clin Epidemiol [Internet]. 2011 Apr [cited 2022 Aug 25];3:133. Available from: https://pubmed.ncbi.nlm.nih.gov/21487452/

25. Perkins J. Python Text Processing with NLTK 2.0 Cookbook. 2010.

26. Joulin A, Grave E, Bojanowski P, Mikolov T. Bag of Tricks for Efficient Text Classification. 2016;

27. François Chollet. Deep Learning with Python. 2017. 384 p.

28. Akiba T, Sano S, Yanase T, Ohta T, Koyama M. Optuna: A Next-generation Hyperparameter Optimization Framework. In: Proceedings of the ACM SIGKDD International Conference on Knowledge Discovery and Data Mining. New York, NY, USA: Association for Computing Machinery; 2019. p. 2623–31.

29. Falkner S, Klein A, Hutter F. BOHB: Robust and Efficient Hyperparameter Optimization at Scale. 2018;

30. Pinker E. Reporting accuracy of rare event classifiers. NPJ Digit Med. 2018 Dec;1(1).

31. Tomašev N, Harris N, Baur S, Mottram A, Glorot X, Rae JW, et al. Use of deep learning to develop continuous-risk models for adverse event prediction from electronic health records. Nat Protoc. 2021 May 5;1–23.

32. Niculescu-Mizil A, Caruana R. Predicting Good Probabilities With Supervised Learning. 2005;

33. Lundberg SM, Lee SI. A Unified Approach to Interpreting Model Predictions. Adv Neural Inf Process Syst [Internet]. 2017 May 22 [cited 2022 Aug 25];2017-December:4766–75. Available from: https://arxiv.org/abs/1705.07874v2

34. Kokhlikyan N, Miglani V, Martin M, Wang E, Alsallakh B, Reynolds J, et al. Captum: A unified and generic model interpretability library for PyTorch. 2020 Sep 16;

35. Watkinson PJ, Pimentel MAF, Clifton DA, Tarassenko L. Manual centile-based early warning scores derived from statistical distributions of observational vital-sign data. Resuscitation. 2018 Aug 1;129:55–60.

36. Dziadzko MA, Novotny PJ, Sloan J, Gajic O, Herasevich V, Mirhaji P, et al. Multicenter derivation and validation of an early warning score for acute respiratory failure or death in the hospital. Crit Care. 2018 Oct 30;22(1):1–12.

37. Malycha J, Farajidavar N, Pimentel MAF, Redfern O, Clifton DA, Tarassenko L, et al. The effect of fractional inspired oxygen concentration on early warning score performance: A database analysis. Resuscitation. 2019 Jun 1;139:192–9.

38. Joynes E. More challenges around sepsis: definitions and diagnosis. J Thorac Dis. 2016;8(11):E1467.

